# Autonomous conversational agents for loneliness, social isolation, depression and anxiety in older people without cognitive impairment: Systematic review and meta-analysis

**DOI:** 10.1101/2025.10.15.25338078

**Authors:** Yuto Satake, Harry Costello, Nimesh Naran, Daiki Ishimaru, Manabu Ikeda, Robert Howard

## Abstract

Loneliness is a major psychological challenge in older adulthood, contributing to increased risks of depression, anxiety, and mortality. Conversational agents—technologies that interact with users via natural language—have emerged as potential tools to support psychological wellbeing in later life. This systematic review and meta-analysis evaluated the effects of autonomous conversational agents, including robotic and non-robotic systems, on loneliness, as well as social isolation, depression and anxiety in older people without cognitive impairment.

Seventeen studies with pre–post intervention data were included. Nine used physically embodied robots and eight employed non-robotic agents such as personal voice assistants, chatbots, or screen-based embodied agents. Due to the limited number of high-quality comparison studies, all meta-analyses were based on within-group pre–post comparisons. Meta-analytic results showed mild to moderate improvements in loneliness (standardized mean changes using change score [SMCC] = 0.350, 95% CI: 0.180-0.520) and depression (SMCC = 0.464, 95% CI: 0.327-0.602), with no study reporting symptom worsening. Subgroup analyses suggested a somewhat greater effect for robots. No study included validated measures of social isolation, and only one assessed anxiety.

These findings indicate that conversational agents—particularly social robots—may offer scalable support for older adults’ mental health, with potential especially for reducing loneliness and depression. Nonetheless, methodological limitations, including lack of blinded outcome assessment, inconsistent reporting and heterogeneous intervention designs, underscore the need for more rigorous research. Advances in large language models may further enhance the responsiveness and relevance of these technologies for supporting psychological wellbeing in aging populations.

## Introduction

Loneliness has been associated with increased risks of mortality and mental health conditions such as depression and anxiety (Holt-Lunstad, 2024; S. L. Lee et al., 2021; Santini et al., 2020). With advancing age, experience of bereavement, declining health and reduced income may contribute to fewer opportunities for social interaction, leading to increased feelings of isolation (Luhmann & Hawkley, 2016). Globally, approximately one in four older people experience social isolation (R. H. Teo et al., 2023), and more than 20% live alone in many countries(Japan Cabinet Office, 2024; United nations, n.d.). In response to these concerns, several governments have implemented formal initiatives, such as the appointment of Ministers for Loneliness in the UK and Japan, and the World Health Organization established a Commission on Social Connection in 2023 (World Health Organization, 2024). These developments underscore the urgent need for scalable and accessible interventions to reduce loneliness and support mental wellbeing in later life.

Against this backdrop, conversational agents—systems that interact with users through natural language—have attracted increasing attention (Döring et al., 2022; J. Z. T. Teo et al., 2025). A wide range of technologies has emerged, including social robots, personal voice assistants (PVAs), chatbots, and screen-based embodied agents. These tools hold potential for alleviating loneliness and fostering emotional connection, especially among socially isolated older adults. Recent advances in large language models (LLMs) have dramatically expanded the capabilities of conversational agents as a flexible and empathetic talking partner (Guo et al., 2023; Welivita & Pu, 2024). Our research group also has initiated a UK–Japan collaborative project to explore the use of LLM-equipped robots for reducing loneliness in older adults (Satake et al., 2025). While limitations such as delayed system responses and constrained conversational quality persist, and ethical concerns—including privacy and misinformation—remain, both domains are the focus of ongoing technological development and interdisciplinary debate (Irfan et al., 2025).

Although information-technology interventions (Balki et al., 2022), social robots (Gasteiger et al., 2021; Pu et al., 2019; Yu et al., 2022) and chatbots (Zhang et al., 2024) have been reviewed in the context of older-adult care, a comprehensive synthesis that spans both robotic and non-robotic autonomous conversational agents is still lacking. This gap complicates the design of next-generation, LLM–enabled agents aimed at reducing loneliness in community-dwelling older adults without dementia. Therefore, we conducted a systematic review and meta-analysis to quantify the effects of autonomous conversational agents on loneliness and, in addition, on other psychological outcomes that might plausibly benefit from such interventions—namely depression, social isolation, and anxiety—in older adults without cognitive impairment.

## Methods

### 1. Study Design

Systematic review and meta-analysis conducted in accordance with the Preferred Reporting Items for Systematic Reviews and Meta-Analyses (PRISMA) 2020 guidelines. The review aims to synthesize the evidence regarding the efficacy of autonomous conversational agents on loneliness, social isolation, depression and anxiety in older adults without cognitive impairment.

### 2. Protocol and Registration

The protocol for this review was prospectively registered in the International Prospective Register of Systematic Reviews (PROSPERO) under the registration number (CRD42024605387) on 31 October 2024.

### 3. Eligibility Criteria

Inclusion criteria: 1) Participants aged 60 years or older without cognitive impairment or dementia. 2) Use of autonomous conversational agents (e.g., social robots, chatbots, embodied conversational agents). 3) Reported validated quantitative measures of loneliness, social isolation, depression, and/or anxiety both before and after the intervention. 4) Any type of intervention study, including randomized controlled trials, non-randomized controlled studies and single-group pre-post designs.

Exclusion criteria: 1) Not peer-reviewed. 2) Published in a language other than English or Japanese. 3) Grey literature, defined as non–peer-reviewed sources such as unpublished reports, theses, and conference abstracts. Peer-reviewed conference papers (particularly from engineering fields) were included if they met all other inclusion criteria.

Although our initial eligibility criterion specified participants aged 60 years or older, we found that some studies targeting older adults also included participants under the age of 60. To accommodate these real-world recruitment practices while maintaining our focus on older populations, we included studies in which the mean participant age was 60 years or older. In cases where essential outcome data were not reported in the published articles, we contacted the authors to obtain the necessary information and included the study whenever possible based on the additional data provided.

### 4. Information Sources and Search Strategy

A comprehensive search was conducted using the following electronic databases: Ovid MEDLINE (ALL), Embase (1974–2024), APA PsycINFO, CINAHL Plus (EBSCOhost), Web of Science, IEEE Xplore, ACM Digital Library, Ichushi-Web, CiNii Research, and the National Diet Library. The search included studies published up to 1 November 2024.

For MEDLINE, Embase, PsycINFO, CINAHL Plus, and Web of Science, we performed title, abstract, and keyword searches. For IEEE Xplore, we conducted title and abstract searches. For the remaining databases, we conducted searches without applying filters.

The search terms included Population terms (e.g. "aged" and "elder*"), Technology terms (e.g. "chatbot*", "robot*" and "conversation* agent"), and Outcome terms (e.g. "lonel*" and "depress*"). Further details of the search strategy are presented in **Supplementary Note**. In addition, relevant review articles were manually screened to identify additional studies.

### 5. Selection Process

Initial citation search and the collection of title/abstract were conducted by YS. Title/abstract and full-text screenings were performed independently by two reviewers. For English-language articles, the screening was conducted independently by YS and NN. For Japanese-language articles, the screening was independently conducted by YS and DI. Discrepancies at any stage were resolved through discussion among four reviewers (HC, YS, DI, and NN).

### 6. Data Collection Process

YS imported all retrieved citations into a citation manager (Zotero), and duplicates were removed. Full texts of potentially relevant studies were assessed in detail. Data extraction was performed by YS using a pre-defined spreadsheet and included study characteristics, participant demographics, intervention details, outcome measures, and pre-post change scores. DI and NN cross-checked the spreadsheet.

### 7. Data Items

The following data were extracted: study design, number of participants per group, participant characteristics, intervention type and duration, outcome measures for loneliness, social isolation, depression, and anxiety and corresponding pre- and post-intervention scores.

### 8. Study Risk of Bias Assessment

Two reviewers (YS and DI) independently assessed the risk of bias of each included study. Although we initially planned to use Version 2 of the Cochrane Risk of Bias tool for randomized controlled trials and the Newcastle-Ottawa Scale (NOS) for non-randomized studies, the number of randomized controlled trials with two-group comparisons was too limited. Therefore, NOS was applied to all included studies regardless of study design. Any discrepancies between reviewers were resolved through discussion among the review team.

### 9. Effect Measures

We had planned to use standardised mean differences to compare intervention and control groups. However, due to the limited number of studies with two-group comparisons, we calculated effect sizes within each intervention group using the standardized mean change using change score (SMCC) approach. The primary effect measure was SMCC based on validated scales of loneliness. Additional outcomes included SMCC values for social isolation, depression and anxiety, where data were available. Since none of the included studies reported pre–post correlations required for SMCC calculations, we assumed a correlation coefficient (r) of 0.5 as a conservative default, following common practice in meta-analytic research.

### 10. Synthesis Methods

We conducted meta-analyses using a random-effects model if three or more eligible studies reported on the same outcome. Meta-analyses were performed using R software with the metafor package. We assessed statistical heterogeneity using the I² statistic and explored sources of heterogeneity where appropriate. To investigate potential sources of heterogeneity, we conducted subgroup analyses based on the type of conversational agent used. Specifically, we compared outcomes from studies employing physically embodied robots (e.g., humanoid social robots) with those using non-robotic conversational agents (e.g., text-based chatbots or screen-based avatars).

In addition, we performed leave-one-out (LOO) analyses and excluded small-sample studies to explore their influence on overall results. As part of the sensitivity analyses related to the assumed correlation coefficient (r), we re-ran all meta-analyses using r = 0.3 and r = 0.7 to evaluate the impact of this assumption on the estimated effect sizes.

### 11. Reporting Bias Assessment

Funnel plots was generated and Egger’s test was performed to assess potential publication bias. If asymmetry was observed, we planned to conduct sensitivity analyses and interpret the results with caution, considering possible small-study effects.

## Results

### 1. Study Selection

A total of 2,540 records were identified through database searches. After removing duplicates, 1,445 unique records remained. Of these, 116 full-text articles were assessed for eligibility, and 17 studies met the inclusion criteria. Sixteen of these studies were included in the meta-analyses. The study selection process is illustrated in the PRISMA 2020 flow diagram (**Figure 1**).

**Figure 1.**
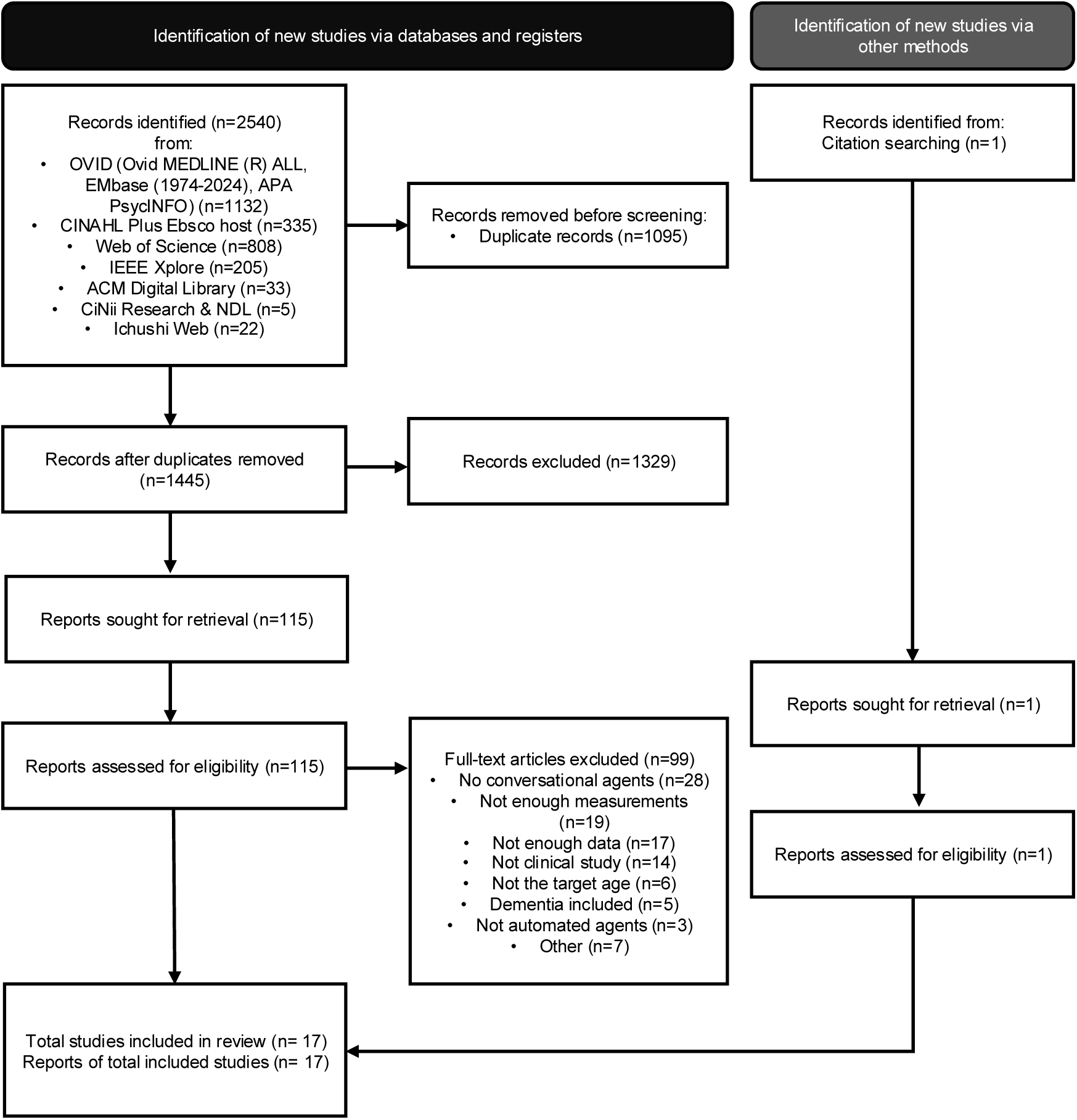
PRISMA 2020 flow diagram.

### 2. Study Characteristics

Study characteristics are summarized in **Table 1**, with further details provided in **Supplementary Table S1**. Sample sizes of the intervention groups ranged from 4 to 291 participants (median = 18; mean = 43.6; SD = 74.1). Participants’ ages ranged from 50 to 98 years, with most studies targeting adults aged 65 years or older. Based on data from 16 of the 17 included studies other than one without reported mean age, the weighted mean age of participants across intervention groups was 78.1 years. Standard deviations were available for 14 studies, and a pooled standard deviation was therefore not calculated. The majority of participants were women, and three studies included only female participants. Of the 17 included studies, ten specifically targeted community-dwelling older adults living alone. Two studies involved community-dwelling older people without restricting for living situation. The remaining five studies were conducted in institutional or assisted living settings, including nursing homes, care homes and retirement villages.

**Table 1.** The summary of included studies.

| Author (Year) | Country | N | Living situation | Diagnosis | Intervention type | Agent description | Duration / Frequency | Outcomes | Summary |
| --- | --- | --- | --- | --- | --- | --- | --- | --- | --- |
| **Yan et al. (2024)** | United States | 15 | Living alone (independent, at home) | Community-dwelling older adults | PVA | Alexa (Amazon Echo: a commercial voice-controlled smart speaker equipped) | 4 weeks; Daily use; $\geq 5$ times/day (instructed); reminders sent on days 7, 14, and 21 if below threshold | Lo | • Significant reduction in loneliness after 4 weeks<br>• Total time spent with Alexa was significantly correlated with loneliness reduction |
| **Wang and Li (2024)** | China | 4 | Nursing home residents | NR; assumed cognitively normal nursing home residents | Chatbot | ChatGPT 3.0 (via headset + screen + researcher-supported dialogue) | 8 weeks; 15–30 min, once / week (8 sessions total) | Lo, De | • No significant in loneliness and depression |
| **Lee et al. (2024)** | United States | 30 | 70% lived alone | Community-dwelling older adults | Robot | Hyodol: doll-shaped socially assistive humanoid robot with speech | 4 months; Daily use; individual usage not strictly prescribed | Lo, De | • Depressive symptoms improved significantly<br>• Loneliness decreased but not significantly |
| **Jones et al.(2024)** | United States | 34 | All participants lived alone in independent living facilities | Community-dwelling older adults; Cognitively normal to mildly impaired (MoCA mean = 23.5) | PVA | Alexa (audio-only, Echo Dot vs. video, Echo Show) | 12 weeks; Daily use; $\geq 30$ min/day (prescribed routine activities) | Lo | • Significant reduction in loneliness<br>• Significant increase in perceived social support |
| **Chou et al.(2024)** | Taiwan | 15 | 47% living alone | Psychiatric outpatients with depression or anxiety disorders | Chatbot | LINE-based health chatbot | 4 weeks; Daily use; 82.5% adherence ($\geq 65$ group) | Lo, De, An | • Significant loneliness reduction<br>• No significant change in depression or anxiety |
| **Lim (2023)** | South Korea | 31 | living alone | Community-dwelling older adults | Robot | PIO: parrot-shaped socially-assistive robot | 6 weeks; 50 min, 2 sessions / week (12 sessions total) | Lo, De | • Significant improvement in loneliness, depression, and cognitive function |
| **Lee et al. (2023)** | South Korea | 169 | 95.3% lived alone; 39% had no contact with children | Community-dwelling older adults | Robot | Hyodol: doll-shaped socially assistive humanoid robot with speech | 3 months (6 months also); Daily use; 24h inactivity triggers alert | De | • Depressive symptoms significantly improved at 3 months but the effect was not maintained at 6 months. |
| **Park and Kim (2022)** | South Korea | 291 | Living alone | Community-dwelling older adults; multiple chronic illnesses common | PVA | NUGU Candle: Commercial Korean AI speaker with voice interaction features | 2 months; Daily use; comparison between frequent ($\geq 5$/week) and intermittent users | Lo, De | • Significant reduction in loneliness and depression<br>• Loneliness reduction more pronounced in frequent users |
| **Papadopoulos et al.(2022)** | United Kingdom and Japan | 12 | Care home residents living in a single room | Older adults without severe dementia, severe mental health issues, or severe frailty | Robot | Pepper: socially assistive humanoid robot with speech. It was culturally personalised (Experimental group). 4 feet tall | 2 weeks; 6 sessions × up to 3 hours each | Lo | • No significant change in loneliness |
| **Kramer et al.(2022)** | Netherlands | 32 | Living alone | Community-dwelling older adults | Screen-based embodied agents | PACO: Two 2D cartoon-style avatars (Herman for nutrition, Ellen for social advice), text-based, non-animated. | 8 weeks; Daily use; self-paced use; median total use 15 hours | Lo | • No significant change in loneliness and eating behavior |
| **Jones et al.(2021)** | United States | 16 | Living alone | Community-dwelling older adults aged 75+ with normal cognitive functioning | PVA | Alexa (Amazon Echo: a commercial voice-controlled smart speaker equipped) | 4 weeks (8 weeks actually but it was not for measurements of loneliness); Daily; Minimum 5 interactions per day during first 4 weeks (mean $\sim 18$/day); voluntary use thereafter | Lo | • Significant reduction in loneliness<br>• Relational greetings to Alexa predicted loneliness reduction |
| **Kargar et al.(2017)** | United States | 7 | Living at an assisted living facility | Older adults with no to moderate depression | Robot | eBEAR: animal-like social robot with facial expression generation and dialogue capabilities | 1 month; 3 sessions per week (up to one hour per session) | De | • Improvement in depression scores in several participants |
| **Broadbent et al.(2016)** | New Zealand | 27 | Living in rest home and nursing home units | Older adults; Mixed cognitive status (35% with AMTS $\leq 6$ suggesting cognitive impairment) | Robot | Guide: Stationary touchscreen robot offering scripted speech interactions. Cafero: Stationary touchscreen robot with similar functions to Guide. | 12 weeks; Robots were available for 14 hours (6AM-8PM) a day at lounge of facilities; no usage requirement enforced | De | • No significant differences in depression, quality of life, or mobility between groups |
| **Ring et al.(2015)** | United States | 7 | Living alone | Older adults without significant depression symptoms (screened with PHQ-2) | Screen-based embodied agents (proactive version) | Animated touchscreen-based avatar using synthetic voice, facial expressions, gestures | 1 week; Daily; proactive version initiated conversation via motion sensor | Lo | • Proactive agent significantly reduced loneliness and increased happiness and comfort compared to passive agent<br>• Longer interaction time correlated with greater loneliness reduction |
| **Broadbent et al.(2014)** | New Zealand | 29 | Independent living apartments in a retirement village | Older adults; cognitive function assessed (AMTS mean = 9.2/10); mostly cognitively intact | Robot | iRobiQ: Small table-top robot with touchscreen. Cafero: Larger robot with touchscreen. | 6 weeks robot condition + 6 weeks no-robot control (cross-over), with an 18-day washout period; Daily; no enforced usage amount | De | • No significant improvements in depression, quality of life, or medication adherence detected |
| **Tanaka et al.(2012)** | Japan | 18 | Living alone | Community-dwelling older female adults without diagnosed dementia (MMSE $\geq 24$) | Robot | Kabochan: doll-shaped socially assistive humanoid robot with speech | 8 weeks; Daily; interactions not strictly standardized | De | • The change of depression scale was not statistically significant.<br>• Cognitive scales showed improvements. |
| **Suzuki et al.(2005)** | Japan | 4 | Living alone | Community-dwelling older female adults without diagnosed dementia | Robot | Ifbot; humanoid robot capable of recognizing speech and displaying emotional expressions | 4 weeks; Daily; encouraged to interact daily | De, Lo | • Some participants showed improvement in depression scores |

N and Diagnosis represent the number of participants and diagnoses in the intervention group. Abbreviations: PVA, personal voice assistant; Lo, loneliness; De, depression; An, anxiety; NR, not reported; MoCA, Montreal Cognitive Assessment; MMSE, Mini-Mental State Examination; PHQ-2, Patient Health Questionnaire-2; AMTS, Abbreviated Mental Test Score.

The most frequently reported additional outcome was quality of life, included in seven studies. Study designs included eight single-arm pre–post studies (including one case series), four non-randomized two-arm studies, and three randomized controlled trials—one of which employed a crossover design and another a delayed intervention model. Three of these were excluded from the effect size calculation due to missing group-level means (Broadbent et al., 2014; Ring et al., 2015; Wang & Li, 2024). Of the 17 studies, 12 assessed loneliness, 11 assessed depression, one assessed anxiety, and none assessed social isolation using validated outcome measures. The Geriatric Depression Scale (GDS) (Yesavage et al., 1982)was used in 10 of the 11 depression studies, and the UCLA Loneliness Scale (in various versions) (Hays & DiMatteo, 1987; D. Russell et al., 1980; D. W. Russell, 1996)was used in 10 of the 12 loneliness studies.

Interventions varied widely in both platform and function. Nine deployed robots, while eight utilized non-robotic conversational agents: four involved PVAs, two used screen-based embodied agents, and two used chatbots. The most commonly used device was the Amazon Echo (three studies) (Jones et al., 2024; Jones V.K. et al., 2021; Yan C. et al., 2024), followed by Hyodol (two studies) (O. E. Lee et al., 2024; Lee O.E.K. et al., 2023). Only one study deployed LLM-based conversational agents (Wang & Li, 2024). All other studies used different devices or applications. Agent functions ranged from conversation-based interaction to features such as riddles, online shopping support, reminders, games, video calling (e.g., Skype), encouragement for physical activity, singing, dancing, dietary suggestions and cognitive training. Intervention duration ranged from three brief sessions to four months, with usage intensity varying from short daily interactions to continuous availability.

### 3. Risk of Bias in Studies

The mean NOS score was 4.1 (SD = 0.3), with a range of 2 to 7. One study was rated as high quality (7–9 points), 10 studies as moderate quality (4–6 points), and 6 studies as low quality (0–3 points) (**Table 2**). Notably, some NOS items—such as "Selection of non-exposed cohort" and "Comparability of cohorts"—were rated as “not applicable” for single-arm designs. Importantly, no study received a star for "Assessment of outcome," reflecting the absence of blinded outcome evaluation. While most studies received stars for "Ascertainment of exposure" and "Was follow-up long enough for outcomes to occur?", the actual quality of implementation in these items varied. Although not formally assessed within the NOS framework, several two-arm studies studies included participants with notably high educational backgrounds in the intervention groups, which is one of the essential selection biases (Lim J., 2023; Papadopoulos et al., 2022; Wang & Li, 2024).

**Table 2.** Risk of bias assessment using the Newcastle-Ottawa Scale.

| Author (Year) | Total Score (max 9 ☆) | Selection (max 4 ☆) |  |  |  | Comparability (max 2 ☆) | Outcome (max 3 ☆) |  |  |
| --- | --- | --- | --- | --- | --- | --- | --- | --- | --- |
|  |  | Representativeness of the exposed cohort | Selection of the non exposed cohort | Ascertainment of exposure | Demonstration that outcome of interest was not present at start of study | Comparability of cohorts on the basis of the design or analysis | Assessment of outcome | Was follow-up long enough for outcomes to occur | Adequacy of follow up of cohorts |
| Yan et al. (2024) | 4 | ☆ | NA | ☆ | ☆ | NA |  | ☆ |  |
| Wang et al. (2024) | 3 |  | ☆ | ☆ |  |  |  | ☆ |  |
| Lee et al. (2024) | 5 | ☆ | NA | ☆ | ☆ | NA |  | ☆ | ☆ |
| Jones et al. (2024) | 4 | ☆ | NA | ☆ | ☆ | NA |  | ☆ |  |
| Chou et al. (2024) | 3 | ☆ | NA | ☆ |  | NA |  | ☆ |  |
| Lim (2023) | 6 | ☆ | ☆ | ☆ | ☆ |  |  | ☆ | ☆ |
| Lee et al. (2023) | 5 | ☆ | NA | ☆ | ☆ | NA |  | ☆ | ☆ |
| Park and Kim (2022) | 5 | ☆ | NA | ☆ | ☆ | NA |  | ☆ | ☆ |
| Papadopoulos et al. (2022) | 4 |  | ☆ | ☆ |  | ☆☆ |  |  |  |
| Kramer et al. (2022) | 4 | ☆ | NA | ☆ | ☆ | NA |  | ☆ |  |
| Jones et al. (2021) | 3 | ☆ | NA | ☆ |  | NA |  | ☆ |  |
| Kargar et al. (2017) | 3 | ☆ | NA | ☆ |  | NA |  | ☆ |  |
| Broadbent et al. (2016) | 2 |  | ☆ |  |  |  |  | ☆ |  |
| Ring et al. (2015) | 5 | ☆ | ☆ | ☆ | ☆ |  |  |  | ☆ |
| Broadbent et al. (2014) | 3 |  | ☆ | ☆ |  |  |  | ☆ |  |
| Tanaka et al. (2012) | 7 | ☆ | ☆ | ☆ | ☆ | ☆ |  | ☆ | ☆ |
| Suzuki et al. (2005) | 4 | ☆ | NA | ☆ | ☆ | NA |  | ☆ |  |

### 4. Results of Individual Studies

Among the included studies, ten assessed loneliness and nine assessed depression using validated pre- and post-intervention measures in the intervention group. The effect sizes and their 95% confidence intervals for each individual study are presented in **Figure 2**. For loneliness, SMCCs ranged from 0.02 to 0.96, and for depression, they ranged from 0.18 to −0.73. Notably, no study showed a negative effect (i.e., worsening of symptoms). Anxiety was evaluated in only one study using a chatbot intervention (Chou et al., 2024), which reported no significant pre–post change in anxiety with a SMCC score of 0.23 (95% CI [0.10-0.56]) suggesting a reduction in anxiety symptoms. Although a study measured a subjective measure of social support (Jones et al., 2024), no studies deployed an objective scale for social isolation.

**Figure 2.**
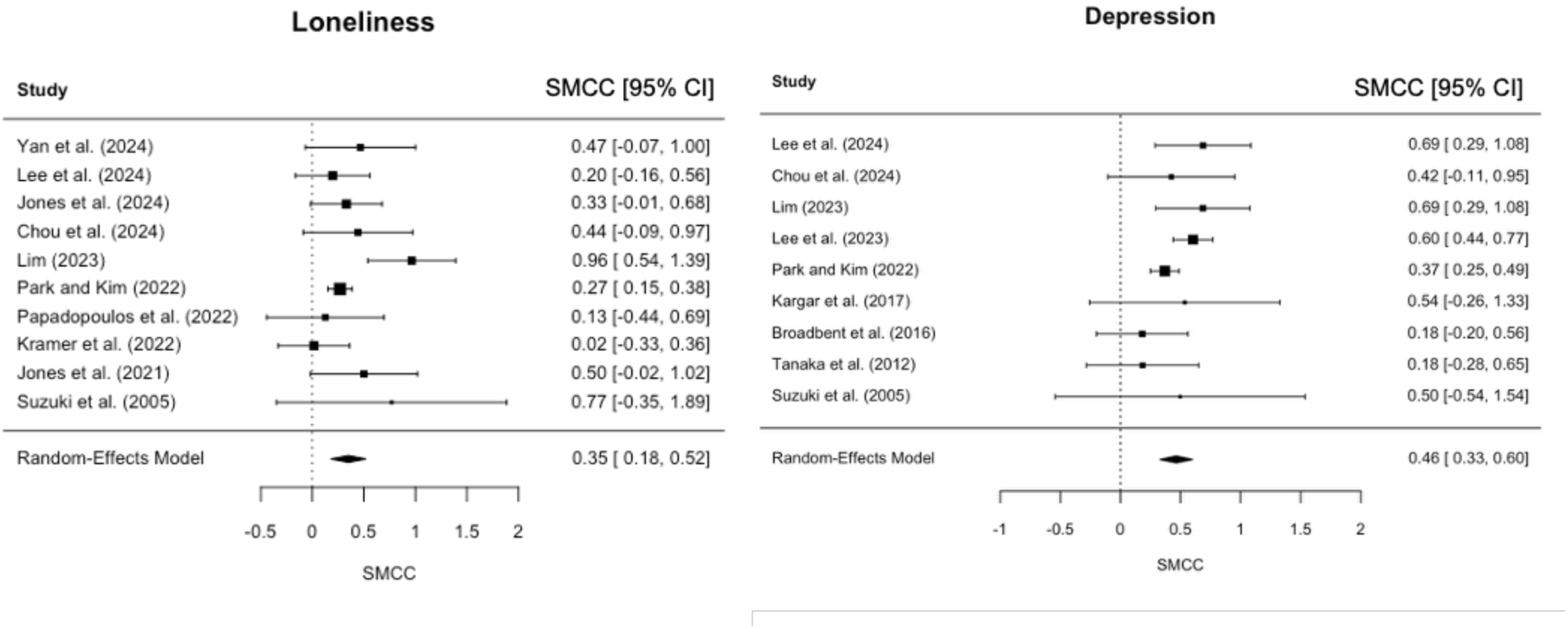
Forest plots of pooled standardized mean change using change score (SMCC) Each study’s point estimate and 95% confidence interval (CI) are shown. The left panel shows results for loneliness, and the right panel shows results for depression. SMCC was calculated by dividing the mean pre–post difference by the standard deviation of the change scores, assuming a pre–post correlation of r = 0.5. Positive values indicate improvements (i.e., reductions in loneliness or depressive symptoms) following the intervention. Pooled effect sizes were estimated using a random-effects model.

### 5. Results of Syntheses

Meta-analyses were conducted for loneliness and depression only based on the number of included papers.

Ten studies were included in the meta-analysis for loneliness. The pooled SMCC was 0.350 (95% CI: 0.180 to 0.520, p < 0.001), indicating a mild to moderate reduction in loneliness following interventions with conversational agents (**Figure 2**). Statistical heterogeneity was moderate (I² = 46.8%, τ² = 0.029), and the heterogeneity test was not statistically significant (Q(9) = 14.82, p = 0.096). Subgroup analyses comparing studies using robotic agents (k = 4) versus non-robotic agents (k = 6) showed a somewhat larger pooled effect in the robotic group (0.479, 95% CI: 0.023 to 0.935, p = 0.039, I² = 65.5%) compared to the non-robotic group (0.274, 95% CI: 0.174 to 0.374, p < 0.001, I² = 0%) (**Figure 3**). These findings suggest a potential advantage for physically embodied agents, although subgroup comparisons should be interpreted with caution due to small sample sizes and study heterogeneity.

**Figure 3.**
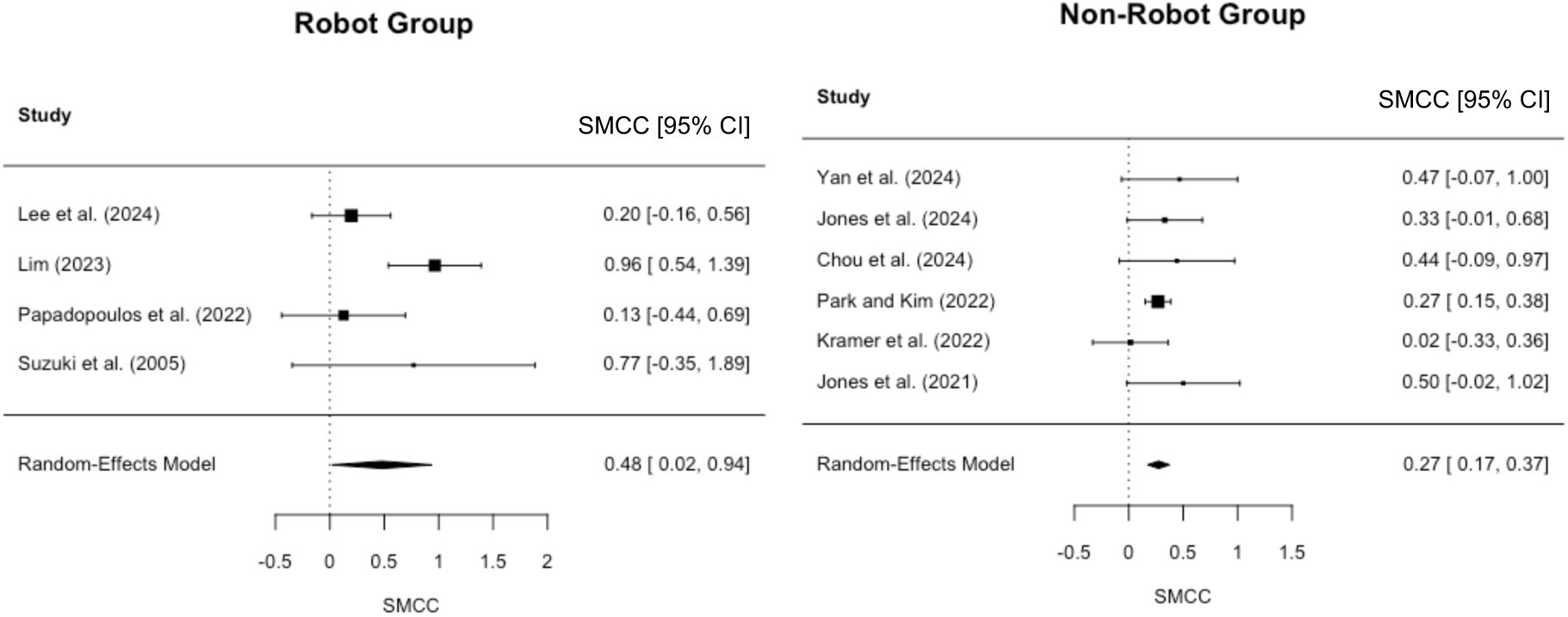
Subgroup analysis of standardized mean change using change score (SMCC) for loneliness. Each study’s point estimate and 95% confidence interval (CI) are shown. The left panel presents results from studies using physically embodied robots, and the right panel includes studies using non-robotic conversational agents. SMCC was calculated by dividing the mean pre–post difference by the standard deviation of the change scores, assuming a pre–post correlation of *r* = 0.5. Positive values indicate reductions in loneliness following the intervention. Pooled effect sizes for each subgroup were estimated using random-effects models.

Nine studies were included in the meta-analysis for depression. The pooled SMCC was 0.464 (95% CI: 0.327 to 0.602, p < 0.001), indicating a mild to moderate reduction in depressive symptoms following the intervention (**Figure 2**). Heterogeneity was low to moderate (I² = 35.9%, τ² = 0.013), and the test for heterogeneity was not statistically significant (Q(8) = 11.14, p = 0.194).

Subgroup analysis was not performed for depression due to the limited number of studies using non-robotic agents (n = 2). LOO analyses were conducted for both loneliness and depression to assess the robustness of the findings. In both cases, exclusion of individual studies did not materially alter the overall pooled effect sizes, indicating that no single study unduly influenced the results (**Supplementary Figure S1)**. Additionally, sensitivity analyses were performed using alternative assumptions for the pre–post correlation used in the SMCC calculation. The primary analysis used r = 0.5, but analyses were also run using r = 0.3 and r = 0.7. The pooled effect sizes remained stable across these values, suggesting that the results were not sensitive to variations in the assumed correlation (**Supplementary Figures S2 and S3**).

### 6. Reporting Biases

To assess potential reporting biases, funnel plots were visually inspected for asymmetry in the meta-analyses of loneliness and depression. As shown in **Supplementary Figure S4**, the distribution of effect sizes appeared generally symmetrical for both outcomes, suggesting a low risk of publication bias. We also conducted Egger’s regression test to statistically assess funnel plot asymmetry. The test indicated no significant evidence of small-study effects for either outcome: loneliness (z = 1.0, p = 0.295) and depression (z = –0.04, p = 0.967). These findings support the visual interpretation that the risk of publication bias was low for both outcomes.

## Discussion

This systematic review and meta-analysis included 17 studies evaluating the effects of autonomous conversational agents on loneliness, depression, social isolation and anxiety in older adults without cognitive impairment. Although several studies included a control group, only six provided sufficient data for group-level comparisons, and all meta-analytic effect sizes were therefore based on pre–post changes within intervention groups. Meta-analytic results indicated small to moderate improvements in loneliness (SMCC = 0.350) and depressive symptoms (SMCC = 0.464), with no evidence of symptom worsening in any study. Subgroup analyses for loneliness suggested that physically embodied social robots may yield greater benefits than non-robotic agents, though the small number of robot-based studies merits caution. Sensitivity analyses confirmed the robustness of findings, and no significant publication bias was detected. These results support the potential utility of conversational agents, particularly social robots, in promoting psychological wellbeing among older adults without cognitive impairment.

This review found that research on conversational agents for cognitively unimpaired older adults has primarily focused on loneliness and depression, with anxiety and social isolation rarely addressed. Only one study assessed anxiety (Chou et al., 2024), and none included validated measures of social isolation. This is somewhat surprising, given that both are well-documented challenges in later life (Holt-Lunstad, 2024) and often examined in conjunction with loneliness in related research fields(Gadbois et al., 2022; Kwok et al., 2024; Lai et al., 2020). The absence of social isolation measures may reflect the dominance of self-reported psychological constructs in this field or a lack of consensus on how to operationalize social connectivity objectively. The consistent use of the UCLA

Loneliness Scales (in various versions) and the GDS suggests a preference for well-validated, standardized tools in this literature. However, inconsistencies in the reported score ranges—where some outcomes deviated from the expected theoretical limits of the scales—were occasionally observed, raising concerns about scoring accuracy or reporting clarity in a few studies.

The findings of this review suggest that conversational agents, including social robots and voice assistants, may help alleviate loneliness and depressive symptoms in older adults. To our knowledge, no previous meta-analysis has specifically focused on conversational agents in this population, and only a limited number of reviews have addressed the topic. For example, recent scoping and systematic reviews have reported potential benefits of PVAs and screen-based embodied agents for loneliness and depression among older adults, although the evidence remains preliminary (Castro Martínez et al., 2025; J. Z. T. Teo et al., 2025). One meta-analysis found a moderate effect of relational agents—including non-conversational robots—on loneliness (Sha et al., 2024). Other reviews have also reported promising effects of social robots on psychological outcomes in older populations (Chen et al., 2018; Yen et al., 2024). By contrast, some reviews have raised concerns about the quality and quantity of available evidence, concluding that the effectiveness of social robots remains uncertain (Pu et al., 2019; Yu et al., 2022). This concern also applies to the present review: none of the 17 included studies implemented blinded outcome assessment, indicating a potential risk of detection bias. Moreover, although several studies included comparison groups, only five were able to be meta-analysed as comparison study, and their methodological quality was limited. Therefore, we opted to analyze within-group changes using a single-arm pre–post design to explore the psychological potential of conversational agents in cognitively unimpaired older adults. In addition, the diversity in intervention duration, intensity, and follow-up periods, along with inconsistencies in reporting dropout rates, highlights the need for greater methodological standardization to improve future comparability and cumulative synthesis.

The subgroup analyses for loneliness conducted in this review suggested that physically embodied social robots may produce somewhat greater effects compared to non-robotic conversational agents. According to Weiss, emotional loneliness arises from a lack of companionship, which offers reassurance of worth and an outlet for intimate self-disclosure (Weiss & Bowlby, 1980). It is well established that humans tend to respond to media such as computers and television as if they were social entities (Nass & Moon, 2000; Reeves & Nass, 1996). However, which features of such agents actually foster a sense of companionship remains unclear. Prior studies have indicated that nonverbal features—such as facial expressions, synchronized gestures and gaze—can enhance the perception of companionship in embodied conversational agents and social robots (Kargar et al., 2017; Ring et al., 2015). Furthermore, in a study using a personal voice assistant, participants with higher baseline loneliness scores were more likely to greet the device as they would a person, and such behaviour predicted subsequent reductions in loneliness (Jones V.K. et al., 2021). Similarly, in a study involving a doll-shaped social robot (Hyodol), participants spontaneously referred to the robot as a “grandchild” or “companion,” suggesting that personification may have played a role in enhancing emotional engagement and therapeutic benefit (O. E. Lee et al., 2024). It is possible that physical embodiment— through gaze, movement, and affective expression—fosters anthropomorphism and, in turn, strengthens companionship, which may help explain why robots were more effective in alleviating loneliness. However, the interventions included in this review were diverse in both form and function, often combining features such as reminders, music playback, and health-related messages. As such, it remains difficult to isolate which specific elements contributed most to the observed effects. Future research should aim to disentangle these components and examine the relative impact of conversational flexibility, physical expression, and other design features (**Supplementary Figure S5**).

This review has several methodological strengths. To our knowledge, it is the first systematic review and meta-analysis to compare the effects of both robotic and non-robotic conversational agents in older adults. We examined a wide range of outcomes—including loneliness, depression, and anxiety—and also collected data on background characteristics such as living situation and cognitive status. Robustness of the findings was assessed through sensitivity analyses and evaluation of publication bias. However, there are several limitations to consider. First, all analyses were based on within-group pre–post changes in the intervention group, which are inherently more susceptible to confounding bias. Moreover, only one study was rated as high quality in the risk of bias assessment, and many studies were judged to have multiple sources of bias. The interventions varied substantially in terms of content, agent type, functionality, and usage frequency, making it difficult to isolate which components were most effective. While studies targeting older people with diagnosed cognitive disorders were excluded, we accepted studies where some participants may have had mild cognitive impairment, provided that the interventions were not designed for cognitively impaired populations. In addition, participant characteristics were not consistent across studies; several samples were skewed toward individuals with higher levels of education or socioeconomic status, which may limit the generalizability of the findings. Finally, most studies did not include long-term follow-up, so the durability of the observed effects remains unclear.

## Conclusion

This systematic review highlights that conversational agents—including both robotic and non-robotic systems—have the potential to support psychological well-being among community-dwelling older adults without cognitive impairment. Although the overall evidence base remains limited and heterogeneous, particularly in terms of study quality and intervention duration, the findings suggest that such technologies may help alleviate loneliness and depressive symptoms. This trend will be supported by the development of LLMs. Further high-quality, long-term studies with rigorous designs are needed to clarify their effectiveness and inform future development and implementation.

## Data Availability

All data analyzed in this study are derived from previously published studies and are fully available within the manuscript and its supplementary materials.

## Acknowledgements

We acknowledge the use of artificial intelligence (AI) tools during the development of this manuscript. OpenAI’s ChatGPT (GPT-4o and o3; https://chat.openai.com) was used between April and May 2025 to assist with idea generation, language refinement, and summarisation of extracted data. The research design, study selection, and screening process were conducted independently by the authors, and all outputs generated by AI were reviewed and revised by the authors for accuracy and clarity. In addition, the illustration of Supplementary Figure S5 was created with the assistance of Gamma (https://gamma.app) in April 2025. The authors confirm that no generative AI tool was used to create novel scientific content or to analyze study data, and all final decisions regarding content, structure, and interpretation were made by the authors.

## Authors’ contributions

YS designed the study, conducted data collection, screening, statistical analysis, and drafted the manuscript. NN screened English-language articles; DI screened Japanese articles and reviewed the risk of bias. HC supervised the study design and analysis. RH supervised the project and manuscript preparation. MI critically reviewed the manuscript and suggested improvements. All authors approved the final version of the manuscript.

## Ethical standards

This study is a systematic review and meta-analysis based on previously published data and did not involve human participants. Therefore, ethical approval was not required.

## Supplementary Note. Search Terms

Citation Search 1/11/2024

### English Databases

- **Ovid search**Filter: TI/AB/Keyword heading

Medline Ovid MEDLINE (R) ALL
Embase (1974-2024)
APA PsycINFO
- **CINAHL Plus Ebsco host** Filter: TI/AB/MW
- **Web of Science** Filter: TI/AB/author keywords (Search Terms)

"aged" OR "elder*" OR "older*" OR "geriatric" OR "senior"
AND
"chat bot*" OR "chat-bot*" OR "chatbot*" OR "robot*" OR "avatar*" OR "large language model" OR "artificial intelligence" OR “digital human” OR “virtual human” OR "conversation* agent" OR "AI agent" OR "virtual agent" OR "digital* agent" OR "voice agent" OR "chat agent" OR "relation* agent" OR "companion* agent" OR "comput* agent" OR "conversation* assistant" OR "AI assistant" OR "virtual assistant" OR "digital* assistant" OR "voice assistant" OR "chat assistant" OR "relation* assistant" OR "companion* assistant" OR "comput* assistant" OR "dialog* system" OR "natural language interface" OR "conversation* interface"
AND
"lonel*" OR "isolat*" OR "social exclusion" OR "social particip*" OR "social connect*" OR "social segregation" OR "social companion*" OR "social relation*" OR "depress*" OR "anxi*"
- **IEEE Xplore** Filter: Document title/AB (Search Terms)

"aged" OR "elderly" OR "older" OR "geriatric" OR "senior"
AND
"chat bot" OR "chatbot" OR "robot" OR "avatar" OR "conversational" OR "AI" OR "language" OR "virtual " OR "digital " OR "voice" OR "chat"
AND
"lonely" OR "loneliness" OR "isolat*" OR "depress*" OR "anxi*"
- **ACM Digital Library** Filter: none (Search Terms)

(Title:("aged" OR "elderly" OR "older" OR "geriatric" OR "senior") OR Abstract:("aged" OR "elderly" OR "older" OR "geriatric" OR "senior"))
AND
(Title:("chat bot" OR "chatbot" OR "robot" OR "avatar" OR "conversational" OR "AI" OR "language" OR "virtual " OR "digital " OR "voice" OR "chat") OR Abstract:("chat bot" OR
"chatbot" OR "robot" OR "avatar" OR "conversational" OR "AI" OR "language" OR "virtual " OR "digital " OR "voice" OR "chat"))
AND
(Title:("lonely" OR "loneliness" OR "isolat*" OR "depress*" OR "anxi*") OR Abstract:("lonely" OR "loneliness" OR "isolat*" OR "depress*" OR "anxi*"))

### Japanese Databases

- **CiNii Research**Filter: 原著論⽂限定
- **the National Diet Library** Filter: 原著論⽂限定 (Search Terms)

"⾼齢*" OR " ⽼⼈" AND
"chat bot*" OR "chat-bot*" OR "chatbot*" OR "チャットボット” OR "robot*" OR “ロボット” OR "アバター" OR "large language model" OR “⼤規模⾔語モデル” OR "artificial intelligence" OR “⼈⼯知能” OR "バーチャル" OR "AI エージェント" OR "AI アシスタント " OR "対話システム"
AND
"孤独*" OR "孤⽴*" OR "うつ" OR "鬱" OR “不安”
- **Ichushi-Web**Filter: ⽇本語限定、原著論⽂限定 (Search Terms)

"⾼齢"/AL or " ⽼⼈"/AL
AND
"chat bot"/AL or "chat-bot"/AL or ("⽣成的⼈⼯知能"/TH or "chatbot"/AL) or ("⽣成的⼈⼯知能"/TH or "チャットボット"/AL) or "robot"/AL or ("ロボット"/TH or "ロボット"/AL) or ("アバター"/TH or "アバター"/AL) or "large language model"/AL or "⼤規模⾔語モデル"/AL or ("⼈⼯知能"/TH or "artificial intelligence"/AL) or ("⼈⼯知能"/TH or "⼈⼯知能"/AL) or "バーチャル"/AL or "AI エージェント"/AL or "AI アシスタント"/AL or "対話システム"/AL
AND
"孤独*"/AL or "孤⽴*"/AL or "うつ"/AL or "鬱"/AL or ("不安"/TH or "不安"/AL)

**Supplementary Figure S1.**
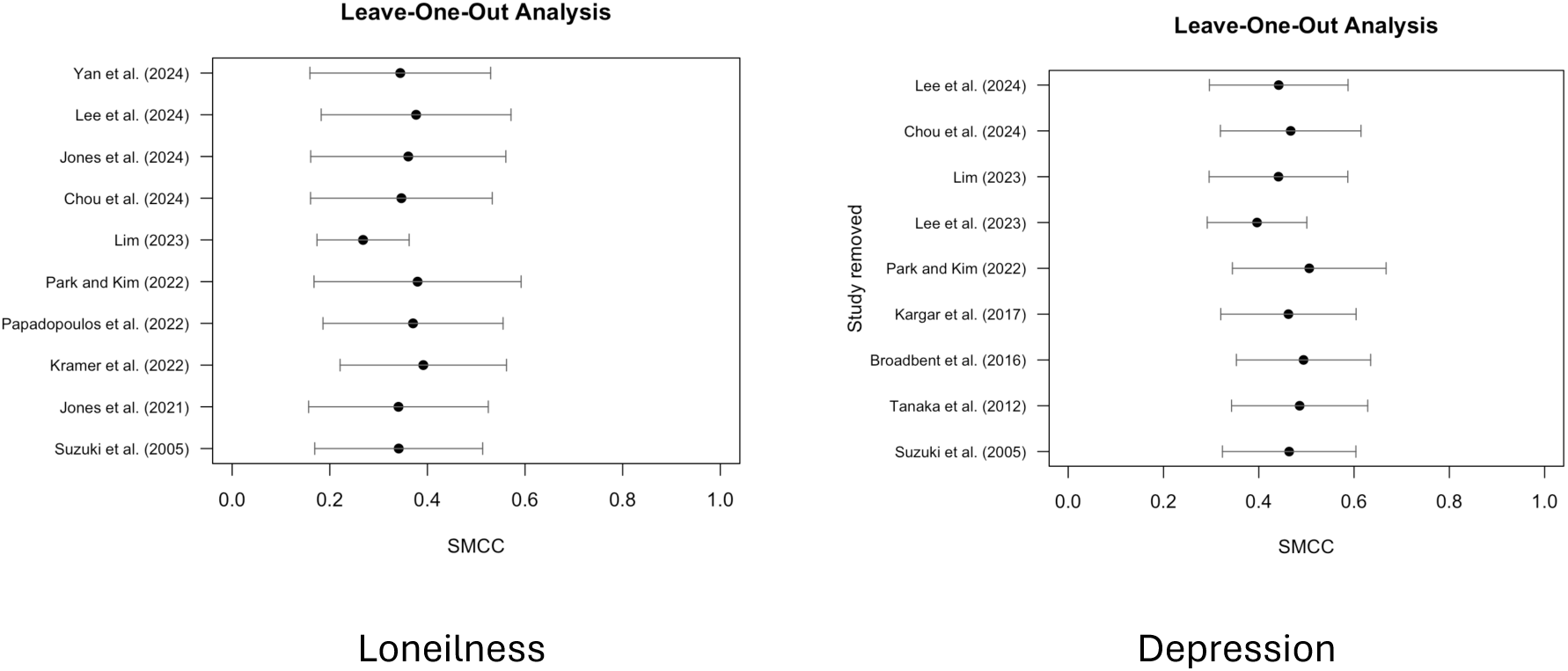
Leave-one-out sensitivity analysis for loneliness and depression These figures show the changes in the pooled standardized mean change with change score (SMCC) for loneliness and depression when each included study is sequentially removed from the meta-analysis.

**Supplementary Figure S2.**
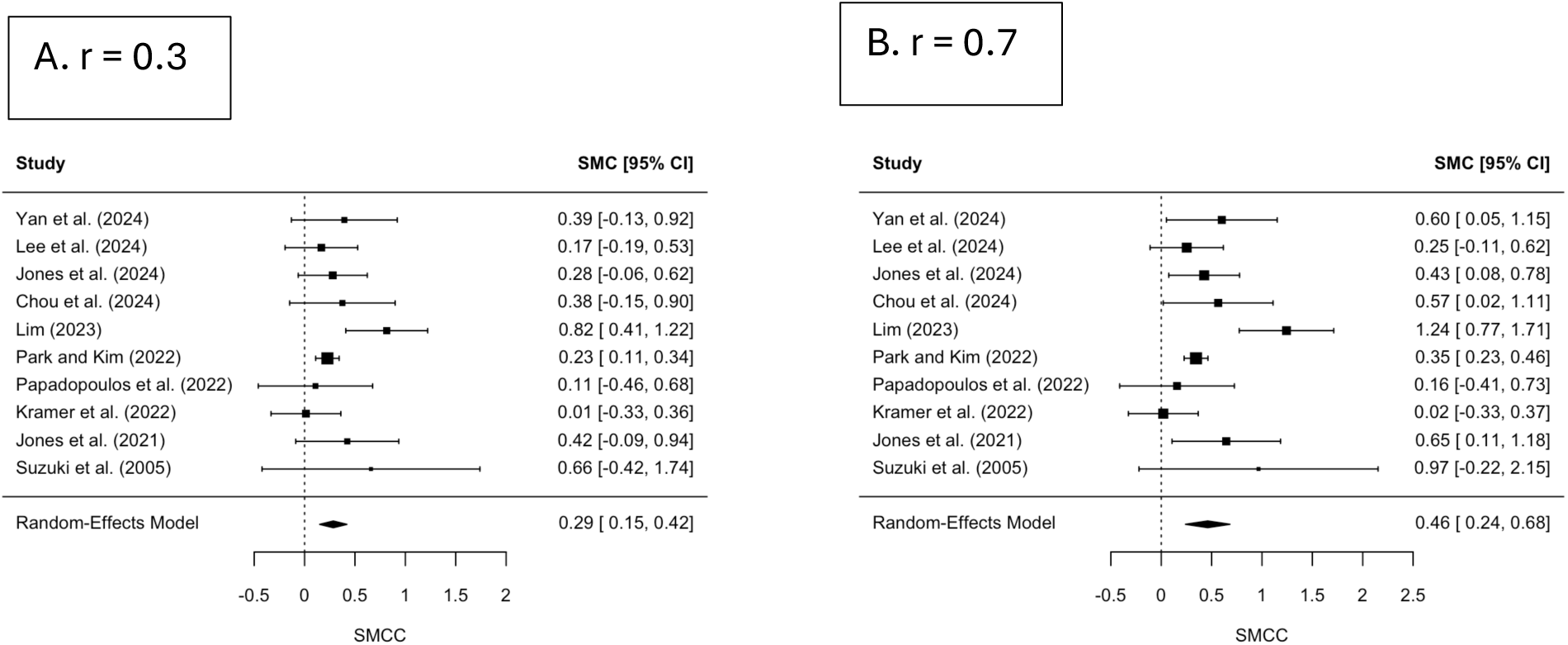
Sensitivity analysis of pooled SMCC for loneliness under different pre–post correlation assumptions The forest plots show the pooled standardized mean change with change score (SMCC) for loneliness calculated using two different assumed values (r=0.3, 0.7) for the correlation between pre- and post-intervention scores.

**Supplementary Figure S3.**
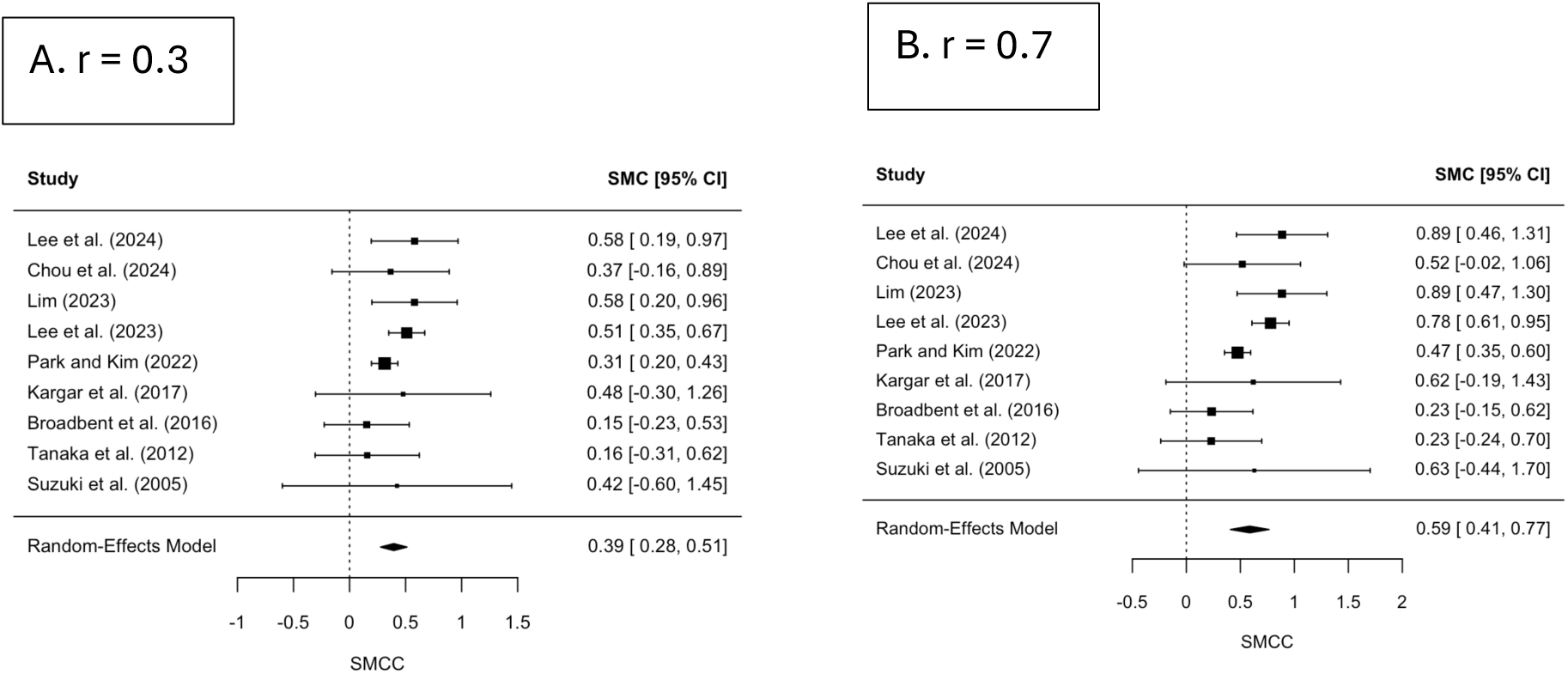
Sensitivity analysis of pooled SMCC for depression under different pre–post correlation assumptions These figures illustrate the robustness of the pooled standardized mean change with change score (SMCC) for depression when varying the assumed correlation between pre- and post-intervention scores.

**Supplementary Figure S4.**
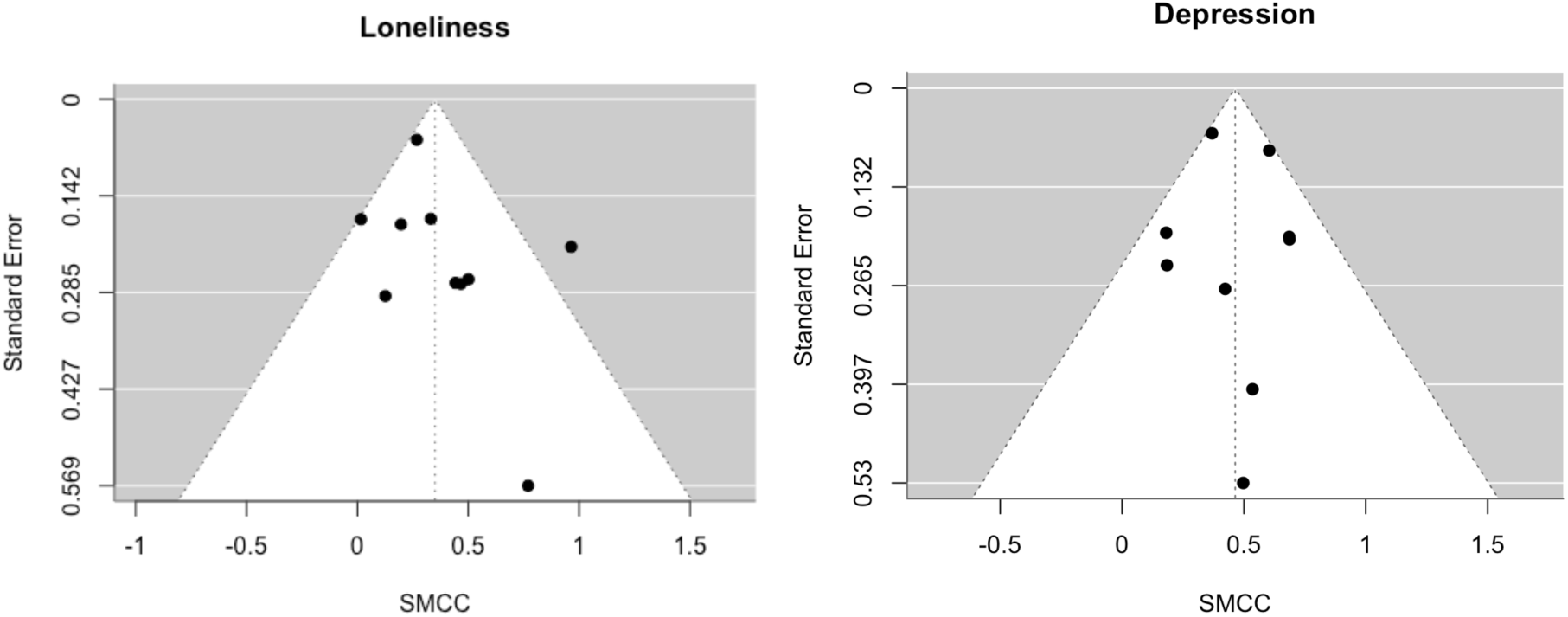
Funnel plot of studies reporting loneliness and depression outcomes

**Supplementary Figure S5.**
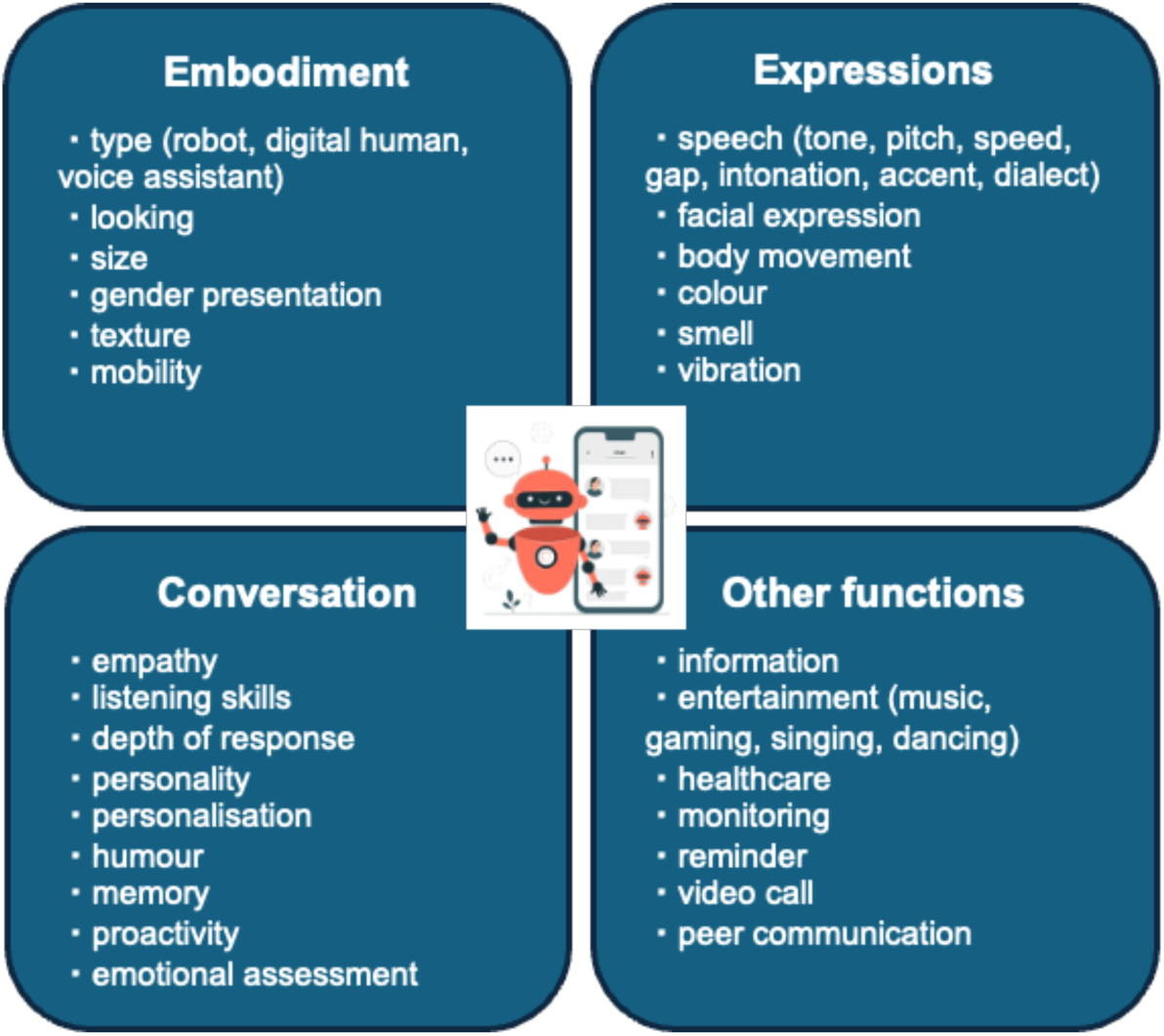
Modifiable design features of conversational agents. Key design domains of conversational agents include embodiment, expressive capabilities, conversational characteristics, and other interactive functions. Each domain encompasses several modifiable factors that can influence user experience, engagement, and intervention effectiveness. These include physical or virtual appearance (e.g., robot, avatar, voice assistant), speech and non-verbal expressions, personality traits and memory, as well as functionalities such as reminders, entertainment, and video calls.

**Supplementary Table S1.**
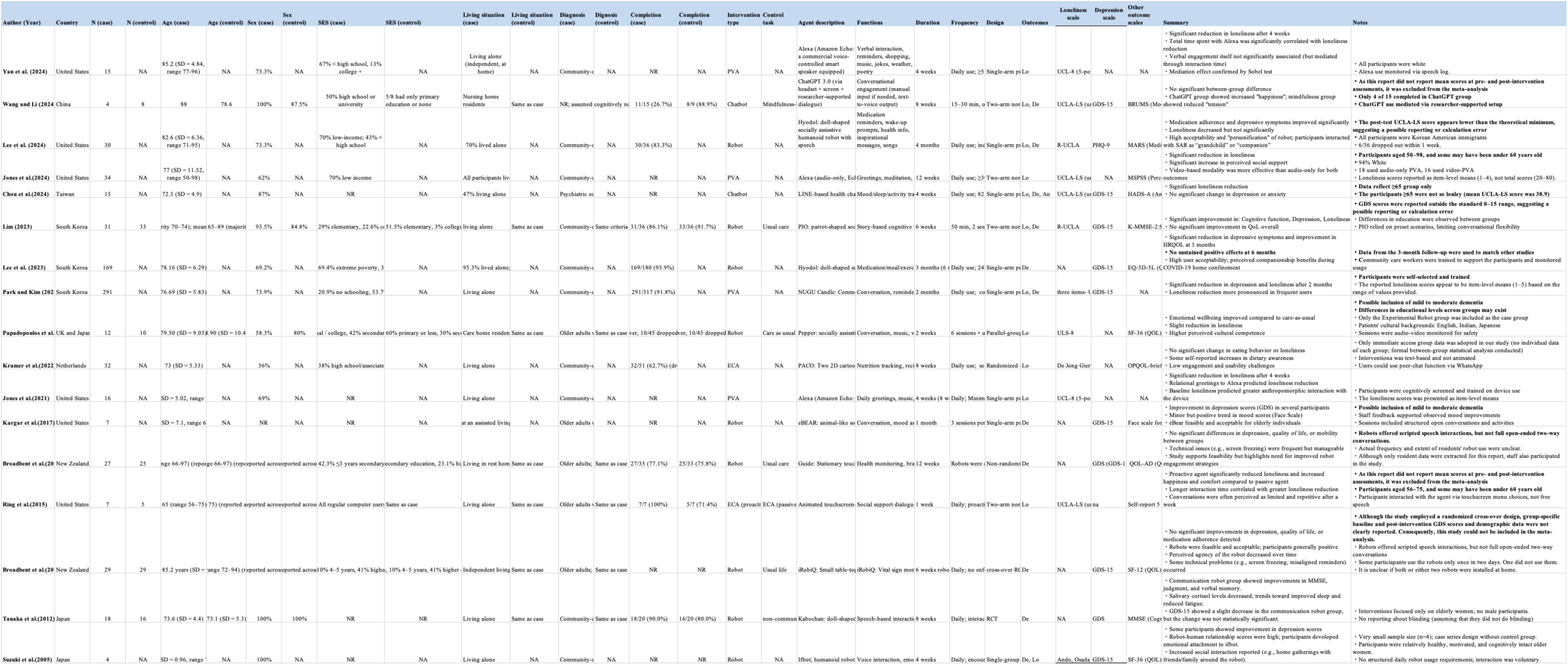
The details of selected articles.

